# SARS-CoV-2 lineage B.1.1.7 is associated with greater disease severity among hospitalised women but not men

**DOI:** 10.1101/2021.06.24.21259107

**Authors:** Oliver T. Stirrup, Florencia A. T. Boshier, Cristina Venturini, José Afonso Guerra-Assunção, Adela Alcolea-Medina, Angela H Becket, Themoula Charalampous, Ana da Silva Filipe, Sharon Glaysher, Tabassum Khan, Raghavendran Kulasegara-Shylini, Beatrix Kele, Irene M. Monahan, Guy Mollett, Matthew Parker, Emanuela Pelosi, Paul Randell, Sunando Roy, Joshua F. Taylor, Sophie J. Weller, Eleri Wilson-Davies, Phillip Wade, Rachel Williams, COG-UK HOCI Variant Substudy consortium, The COVID-19 Genomics UK (COG-UK) consortium, Andrew J. Copas, Teresa Cutino-Moguel, Nick Freemantle, Andrew C. Hayward, Alison Holmes, Joseph Hughes, Tabitha W. Mahungu, Gaia Nebbia, David G. Partridge, Cassie F. Pope, James R. Price, Samuel C. Robson, Kordo Saeed, Thushan I. de Silva, Luke B. Snell, Emma C. Thomson, Adam A. Witney, Judith Breuer

## Abstract

**Background:** Severe acute respiratory syndrome coronavirus 2 (SARS-CoV-2) lineage B.1.1.7 has been associated with an increased rate of transmission and disease severity among subjects testing positive in the community. Its impact on hospitalised patients is less well documented.

**Methods:** We collected viral sequences and clinical data of patients admitted with SARS-CoV-2 and hospital-onset COVID-19 infections (HOCIs), sampled 16/11/2020 - 10/01/2021, from eight hospitals participating in the COG-UK-HOCI study. Associations between the variant and the outcomes of all-cause mortality and intensive therapy unit (ITU) admission were evaluated using mixed effects Cox models adjusted by age, sex, comorbidities, care home residence, pregnancy and ethnicity.

**Results:** Sequences were obtained from 2341 inpatients (HOCI cases = 786) and analysis of clinical outcomes was carried out in 2147 inpatients with all data available. The hazard ratio (HR) for mortality of B.1.1.7 compared to other lineages was 1.01 (95% CI 0.79-1.28, P=0.94) and for ITU admission was 1.01 (95% CI 0.75-1.37, P=0.96). Analysis of sex-specific effects of B.1.1.7 identified increased risk of mortality (HR 1.30, 95% CI 0.95-1.78) and ITU admission (HR 1.82, 95% CI 1.15-2.90) in females infected with the variant but not males (mortality HR 0.82, 95% CI 0.61-1.10; ITU HR 0.74, 95% CI 0.52-1.04).

**Conclusions:** In common with smaller studies of patients hospitalised with SARS-CoV-2 we did not find an overall increase in mortality or ITU admission associated with B.1.1.7 compared to other lineages. However, women with B.1.1.7 may be at an increased risk of admission to intensive care and at modestly increased risk of mortality.

## Introduction

The emergence of severe acute respiratory syndrome coronavirus 2 (SARS-CoV-2) lineage B.1.1.7 in South East England has been found to be associated with an estimated 70% increased rate of community transmission compared with previously circulating variants ^1–3^. Lineage B.1.1.7 is now the dominant lineage in the UK. It has also been detected in over 120 countries outside the UK^4^.

Lineage B.1.1.7 has acquired an unusually large number of mutations and deletions in a short period of time ^1–3^; specifically 14 non-synonymous single nucleotide polymorphisms (SNPs) and 3 amino acid deletions, with 8 of these 17 amino acid changes occurring in the spike protein, responsible for receptor binding and a major immunogenic target. At least three of the spike protein changes are associated with *in vitro* biological changes. A tyrosine substitution at position 501 in the spike protein receptor binding domain has been shown to increase binding to the ACE2 receptor, while deletion of spike protein amino acids 69/70 reduces antibody neutralisation by convalescent sera ^5,6^. The potential that so many mutations might change B.1.1.7 virulence has been examined epidemiologically using data largely from community-collected samples^7^. However, there are few data on the impact of B.1.1.7 infection as compared with other variants on disease outcomes in hospitalized patients.

We investigated the potential associations between the B.1.1.7 variant and the outcomes of mortality and intensive therapy unit (ITU) admission both in patients admitted with COVID-19 and hospital onset COVID-19 infections (HOCIs) in the COG-UK-HOCI study. The main objective was to estimate the overall effect of the variant on each of these outcomes, and we also evaluated whether the impact of the variant differed according to patient characteristics.

## Methods

### Sequence and patient meta-data

Data were collected from five NHS hospitals within London and three outside. The first SARS-CoV-2 positive sample from all inpatients tested through hospital laboratories between 16^th^ November 2020 and 10^th^ January 2021 was sequenced. In addition metadata were collected on patient age, sex (as binary M/F), co-morbidities as identified by the COVID-19 Greenbook^8^ (including obesity with BMI ≥35 kg/m^2^), care home residence, pregnancy, ethnicity, date of hospital admission, ward location and first SARS-CoV-2 positive test for all samples plus dates of admission to the ITU and all-cause death where these events occurred. Ethical approval for the HOCI study is provided by REC 20/EE/0118.

Inpatients were classified as those admitted with SARS-CoV-2 plus cases which were identified after admission, with the latter termed HOCI cases and subdivided into indeterminate healthcare-associated infections (HCAIs) diagnosed 3-7 days after admission and probable/definite HCAIs diagnosed ≥8 days post-admission^9^. The primary outcomes for analysis were the events of death and of ITU admission. Events were included in the analysis within 28 days of hospital admission for those admitted with COVID-19 and within 28 days of diagnosis for HOCI cases.

### SARS-CoV-2 sequencing

Samples were sequenced by Nanopore or Illumina methods as part of the COG-UK consortium. To maximise success 4/8 labs sequenced only those samples with qPCR cycle thresholds (ct) values of ≤32 or equivalent. Sequences were assigned to lineages using COG-UK Pangolin^10^.

### Statistical analysis

Only patients with admission to hospital and HOCIs were included in the statistical analysis of the clinical outcomes of mortality and ITU admission. Mortality and ITU admission were modelled as time-to-event outcomes, from time of hospital admission for those admitted with COVID-19 and from time of diagnosis for HOCI cases, censored at 28 days. Analyses of ITU admission were also censored at patient death. Both outcomes were censored at date of data collection for these variables for each site (between 15^th^ January and 22^nd^ February 2021). Mixed effects Cox models were used with adjustment for sex, patient age (using 5-knot restricted cubic spline), number of comorbidities (none, one, two, ≥three), care home residence, pregnancy, ethnicity (White, Black, Asian, mixed or other) and sample week with separate parameters for London sites and for other sites grouped using the R package coxme v2.2-16^11^. A 5-knot restricted cubic spline^12^ was used for patient age in all analyses to allow flexibility in modelling the relationship with each outcome whilst maintaining a consistent model structure. Random intercept terms were included to reflect clustering of outcomes within hospitals and weekly periods nested within hospitals. Cox models were stratified by HOCI status (allowing for different baseline hazard functions in patients admitted with COVID-19 vs HOCI groups).

Outcomes were analysed on a complete case basis with regards to patient characteristics. This decision was based on the availability of complete data for >90% of patients and the fact that Cox regression gives asymptotically unbiased estimates of an association of interest as long as the missingness is not dependent on both outcome (i.e. death or ITU admission) and exposure (B.1.1.7 status)^13^,^14^. The variable of obesity was analysed as ‘morbid obesity’ vs ‘no record of morbid obesity’ on examination of case notes, and was included in statistical models within the ordinal comorbidities variable.

The primary aim of the analysis was to estimate the overall association between the B.1.1.7 *vs* non-B.1.1.7 strain and the risk of each of the outcomes considered. Exploratory secondary analyses also evaluated interactions between B.1.1.7 status and patient characteristics in estimating the effect on each outcome. Analyses were conducted in R version 4.0.2, using tidyverse collection of packages with all plots generated using ggplot2 and survminer ^15– 18^.

## Results

### Study dataset

Between November 16^th^ 2020 and January 10^th^ 2021 SARS-CoV-2 RNA positive upper respiratory tract samples from 2341 inpatients were sequenced from the 8 participating sites (Table 1 and Supplementary Figure 1). Analysis of clinical outcomes was carried out in 2147 inpatients with all data available. The prevalence of lineage B.1.1.7 was highest in London and Hampshire (South of England), but substantially increased at all sites over the study period (Figure S2).

**Table 1.** Proportion of SARS-CoV-2 due to lineage B.1.1.7 for all inpatient sequenced samples according to patient characteristics

|  | Lineage B.1.1.7<br>(n=1107) | Not lineage B.1.1.7<br>(n=1234) | Total (n=2341) |
| --- | --- | --- | --- |
| **Age Group** |  |  |  |
| 0-11 | 15 (62.5) | 9 (37.5) | 24 (100) |
| 12-24 | 20 (58.8) | 14 (41.2) | 34 (100) |
| 25-34 | 61 (62.9) | 36 (37.1) | 97 (100) |
| 35-49 | 159 (56.2) | 124 (43.8) | 283 (100) |
| 50-69 | 371 (53.2) | 326 (46.8) | 697 (100) |
| 70-79 | 208 (42.2) | 285 (57.8) | 493 (100) |
| 80+ | 273 (38.3) | 440 (61.7) | 713 (100) |
| **Sex** |  |  |  |
| Female | 534 (46.1) | 624 (53.9) | 1158 (100) |
| Male | 573 (48.4) | 610 (51.6) | 1183 (100) |
| **Sample week starting:** |  |  |  |
| 16/11/2020 | 15 (8.4) | 164 (91.6) | 179 (100) |
| 23/11/2020 | 26 (11.6) | 198 (88.4) | 224 (100) |
| 30/11/2020 | 59 (24.2) | 185 (75.8) | 244 (100) |
| 07/12/2020 | 55 (26.8) | 150 (73.2) | 205 (100) |
| 14/12/2020 | 138 (43.8) | 177 (56.2) | 315 (100) |
| 21/12/2020 | 220 (54.6) | 183 (45.4) | 403 (100) |
| 28/12/2020 | 361 (75.2) | 119 (24.8) | 480 (100) |
| 04/01/2021 | 233 (80.1) | 58 (19.9) | 291 (100) |
| **Patient Class** |  |  |  |
| HCW | 7 (36.8) | 12 (63.2) | 19 (100) |
| CAI* | 847 (55.1) | 689 (44.9) | 1536 (100) |
| Indeterminate HCAI† | 54 (25.4) | 159 (74.6) | 213 (100) |
| Probable/definite HCAI‡ | 199 (34.7) | 374 (65.3) | 573 (100) |
| **Region** |  |  |  |
| Glasgow | 91 (31.6) | 197 (68.4) | 288 (100) |
| Hampshire | 74 (60.2) | 49 (39.8) | 123 (100) |
| London | 871 (65.7) | 455 (34.3) | 1326 (100) |
| South Yorkshire | 71 (11.8) | 533 (88.2) | 604 (100) |
| **Ethnicity** |  |  |  |
| White | 540 (39.4) | 829 (60.6) | 1369 (100) |
| Black | 174 (53.4) | 152 (46.6) | 326 (100) |
| Asian | 118 (63.1) | 69 (36.9) | 187 (100) |
| Mixed or other | 186 (67.1) | 91 (32.9) | 277 (100) |
| Unknown | 89 (48.9) | 93 (51.1) | 182 (100) |
| **Patient characteristics** |  |  |  |
| Obese (BMI>=35) | 122 (51) [N=1107] | 117 (49) [N=1234] | 239 (100) [N=2341] |
| Pregnant | 25 (55.6) [N=1103] | 20 (44.4) [N=1234] | 45 (100) [N=2337] |
| Care home resident | 45 (36.3) [N=1107] | 79 (63.7) [N=1232] | 124 (100) [N=2339] |
| **Comorbidities** |  |  |  |
| None | 337 (56.6) | 258 (43.4) | 595 (100) |
| One | 307 (47.3) | 342 (52.7) | 649 (100) |
| Two | 261 (45.9) | 308 (54.1) | 569 (100) |
| Three or more | 202 (38.4) | 324 (61.6) | 526 (100) |
| Not recorded | NA (NA) | 2 (100) | 2 (100) |
| **Died within 28d** | 217 (41.2) [N=1106] | 310 (58.8) [N=1230] | 527 (100) [N=2336] |
| **Admitted to ITU within 28d<sup>‡</sup>** | 220 (65.3) [N=1081] | 117 (34.7) [N=1214] | 337 (100) [N=2295] |

Data shown as *n* (%), with [N] with available data shown where missing values possible.
\*Diagnosed at or ≤2 days from admission. †Diagnosed 3-7 days from admission. ‡Diagnosed ≥8 days from admission. <sup>‡</sup>Excluding patients admitted to ITU prior to SARS-CoV-2 diagnosis. CAI, community-acquired infection; HCAI, healthcare-associated infection; HCW, healthcare worker; ITU, intensive therapy unit.

### Mortality outcome

Death within 28 days was reported in 527 (22.5%) of the 2341 patients. Death was recorded as having occurred following discharge with date of death missing in 5, and these patients have been excluded from analyses. Death within 28 days was recorded in 494/2147 of the patients with all data available, with full 28 days of follow-up in 939/1653 of the other patients. On mixed effects multivariable Cox regression, the overall HR for mortality of lineage B.1.1.7 was 1.01 (95% CI 0.79-1.28, P=0.94) (Table 2). Male sex was found to be a substantial risk factor for mortality (hazard ratio (HR) 1.46 vs female, 1.22-1.75; P<0.001) and age was also strongly associated with the risk of death (Figure 1). The risk of death was higher in care home residents (HR 1.39, 95% CI 1.02 to 1.90, P=0.04) and those with one or more significant comorbidities (HR 1.78 (1.26-2.52) for one comorbidity, 2.03 (1.43-2.88) for two and 2.89 (2.04-4.08) for ≥three *vs* none; P<0.001). Those with ethnicity other than White were estimated to be at higher risk of death, but ethnicity was not a statistically significant predictor when evaluated over all categories (P=0.36). No pregnant women died and so this variable was dropped from the model as a perfect predictor.

**Table 2.** Results of mixed effect Cox regression models for death and intensive therapy unit admission, shown as hazard ratio (95% CI) [P-value]. Models were also adjusted by age using natural cubic splines (as shown in Figure 1).

|  | Outcome: death |  | Outcome: ITU admission |  |
| --- | --- | --- | --- | --- |
| variable | Overall effect | Interaction with sex | Overall effect | Interaction with sex |
| **Var.: B.1.1.7** | 1.01 (0.79 to 1.28)<br>[0.943] |  | 1.01 (0.75 to 1.37)<br>[0.939] |  |
| **Var.: B.1.1.7 in males** |  | 0.82 (0.61 to 1.1)<br>[0.177] |  | 0.74 (0.52 to 1.04)<br>[0.086] |
| **Var.: B.1.1.7 in females** |  | 1.3 (0.95 to 1.78)<br>[0.096] |  | 1.82 (1.15 to 2.9) [0.011] |
| **Sex: male** | 1.46 (1.22 to 1.75)<br>[P<0.001] | 1.77 (1.4 to 2.25) [0] | 1.33 (1.05 to 1.68)<br>[0.017] | 2.42 (1.58 to 3.71) [0] |
| **Ethnicity** | [0.364] | [0.316] | [0.088] | [0.038] |
| **White** | 1.0 (reference) | 1.0 (reference) | 1.0 (reference) | 1.0 (reference) |
| **Asian** | 1.29 (0.96 to 1.73) | 1.31 (0.98 to 1.76) | 1.41 (1.02 to 1.94) | 1.46 (1.06 to 2.01) |
| **Black** | 1.04 (0.68 to 1.58) | 1.06 (0.7 to 1.62) | 1.42 (0.96 to 2.11) | 1.5 (1.01 to 2.23) |
| **Mixed or other** | 1.17 (0.84 to 1.64) | 1.18 (0.84 to 1.66) | 1.41 (1 to 1.99) | 1.49 (1.05 to 2.1) |
| **Care home resident** | 1.39 (1.02 to 1.9)<br>[0.037] | 1.4 (1.03 to 1.92)<br>[0.034] | 0.56 (0.2 to 1.52)<br>[0.254] | 0.57 (0.21 to 1.55)<br>[0.269] |
| **Comorbidities** | [P<0.001] | [P<0.001] | [0.034] | [0.031] |
| **None** | 1.0 (reference) | 1.0 (reference) | 1.0 (reference) | 1.0 (reference) |
| **One** | 1.78 (1.26 to 2.52) | 1.77 (1.26 to 2.5) | 1.25 (0.92 to 1.71) | 1.25 (0.91 to 1.7) |
| **Two** | 2.03 (1.43 to 2.88) | 2.03 (1.43 to 2.87) | 1.24 (0.89 to 1.74) | 1.24 (0.89 to 1.74) |
| **Three or more** | 2.89 (2.04 to 4.08) | 2.89 (2.04 to 4.08) | 0.79 (0.54 to 1.15) | 0.78 (0.53 to 1.14) |
| **Pregnant** |  |  | 0.13 (0.02 to 0.98)<br>[0.048] | 0.13 (0.02 to 0.92)<br>[0.042] |
| **By sample week, London** | [0.273] | [0.22] | [0.007] | [0.013] |
| 16/11/2020 | 0.34 (0.15 to 0.76) | 0.32 (0.14 to 0.72) | 0.58 (0.3 to 1.11) | 0.59 (0.3 to 1.13) |
| 23/11/2020 | 0.94 (0.56 to 1.57) | 0.89 (0.53 to 1.49) | 0.57 (0.31 to 1.03) | 0.57 (0.31 to 1.04) |
| 30/11/2020 | 0.67 (0.4 to 1.12) | 0.64 (0.38 to 1.08) | 0.33 (0.16 to 0.67) | 0.34 (0.17 to 0.7) |
| 07/12/2020 | 0.95 (0.52 to 1.74) | 0.91 (0.5 to 1.67) | 0.79 (0.41 to 1.52) | 0.82 (0.42 to 1.59) |
| 14/12/2020 | 0.82 (0.51 to 1.31) | 0.81 (0.51 to 1.29) | 1.33 (0.85 to 2.08) | 1.3 (0.83 to 2.05) |
| 21/12/2020 | 0.9 (0.61 to 1.33) | 0.89 (0.6 to 1.31) | 1.04 (0.7 to 1.54) | 1.05 (0.7 to 1.57) |
| 28/12/2020 | 1.0 (reference) | 1.0 (reference) | 1.0 (reference) | 1.0 (reference) |
| 04/01/2021 | 0.87 (0.51 to 1.51) | 0.87 (0.5 to 1.5) | 1.11 (0.67 to 1.83) | 1.08 (0.65 to 1.8) |
| **By sample week,**<br>**Elsewhere** | [0.102] | [0.118] | [0.621] | [0.612] |
| 16/11/2020 | 1.07 (0.52 to 2.19) | 1.06 (0.52 to 2.18) | 0.55 (0.1 to 2.91) | 0.56 (0.1 to 3.03) |
| 23/11/2020 | 0.8 (0.38 to 1.67) | 0.79 (0.38 to 1.65) | 0.56 (0.11 to 2.87) | 0.58 (0.11 to 3.02) |
| 30/11/2020 | 1.14 (0.57 to 2.3) | 1.13 (0.56 to 2.28) | 0.66 (0.13 to 3.32) | 0.7 (0.14 to 3.55) |
| 07/12/2020 | 0.91 (0.45 to 1.86) | 0.88 (0.43 to 1.79) | 0.45 (0.08 to 2.61) | 0.48 (0.08 to 2.81) |
| 14/12/2020 | 0.74 (0.37 to 1.47) | 0.73 (0.37 to 1.45) | 0.38 (0.07 to 2.06) | 0.4 (0.07 to 2.2) |
| 21/12/2020 | 1.01 (0.51 to 2) | 0.98 (0.5 to 1.95) | 0.81 (0.17 to 3.82) | 0.86 (0.18 to 4.09) |
| 28/12/2020 | 1.27 (0.63 to 2.53) | 1.24 (0.62 to 2.48) | 1.15 (0.24 to 5.42) | 1.22 (0.25 to 5.84) |
| 04/01/2021 | 1.6 (0.81 to 3.15) | 1.54 (0.78 to 3.04) | 0.74 (0.15 to 3.55) | 0.74 (0.15 to 3.59) |

Var., viral variant. P-values are reported from univariate and multivariate Wald tests.

**Figure 1.**
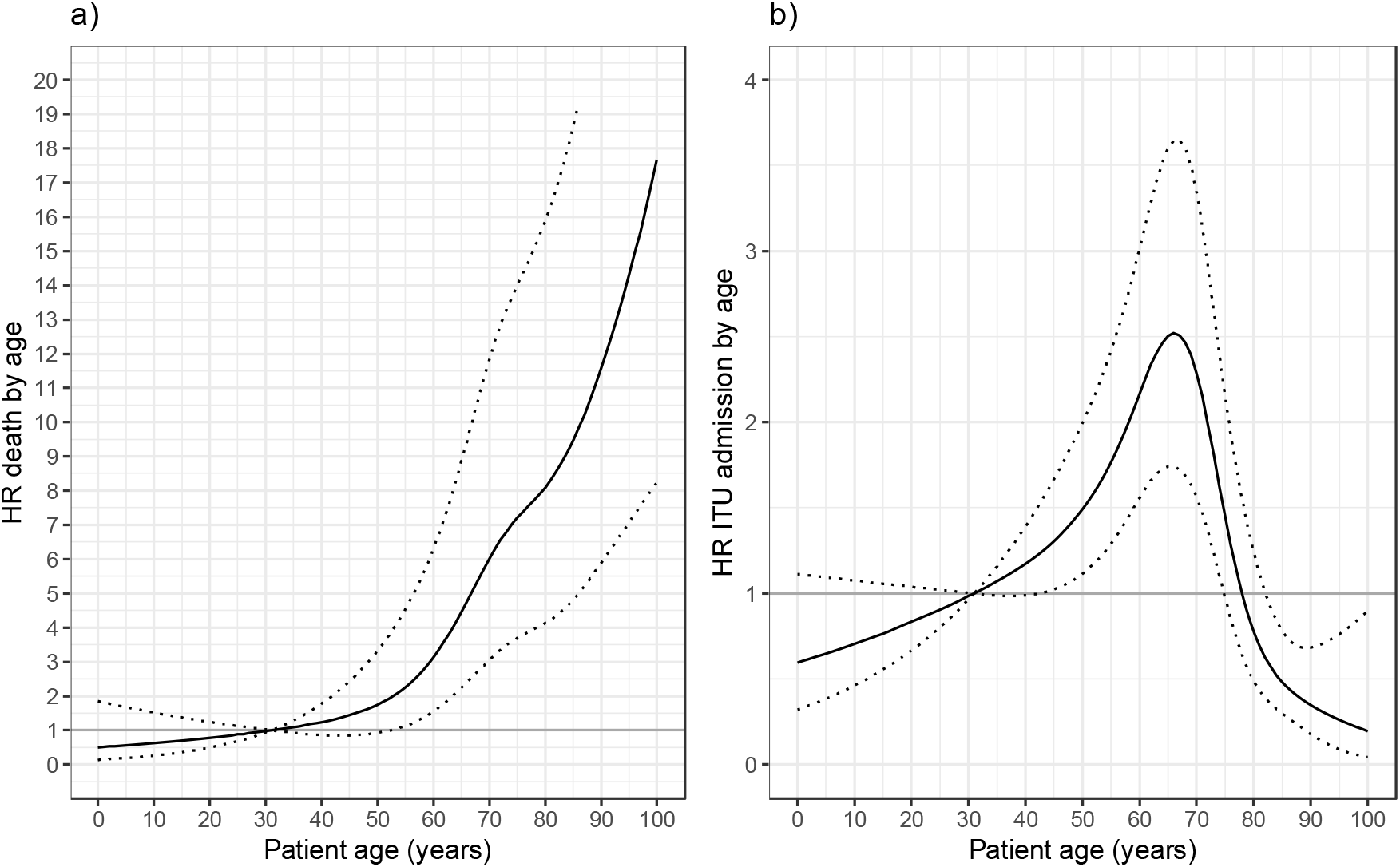
Plots of estimated hazard ratio (HR) for (a) death and (b) intensive treatment unit admission in relation to age for mixed effects Cox regression models with single overall effect of B.1.1.7 variant. Following from the parameterisation of the model, HRs are shown relative to hazard at age of 31 years.

The addition of an interaction term between B.1.1.7 status and patient sex for the effect on mortality led to an improvement in model fit (P=0.01 interaction test, P=0.04 lineage B.1.1.7 effects by sex vs no B.1.1.7 effect, on likelihood ratio tests (LRT)). The estimated HR for mortality of lineage B.1.1.7 vs non-B.1.1.7 was 0.82 (95% CI, 0.61-1.10) in male patients and 1.30 (95% CI, 0.95-1.78) in female patients. No improvement to model fit was provided by the addition of an interaction between B.1.1.7 status and patient age (P=0.48, LRT with 4 d.f.), ethnicity (P=0.67, LRT with 3 d.f.) or comorbidity category (P=0.33, LRT with 3 d.f.).

A statistically significant interaction was found between the effect of B.1.1.7 and care home residence (P=0.03, LRT with 1 d.f.), with those care home residents with B.1.1.7 infection estimated to be at lower risk of death (HR 0.52, 95% CI 0.27 - 1.02) with a non-significant increase in the risk for death associated with B.1.1.7 for non-care home residents (1.09, 95% CI 0.85-1.41). We attempted to fit a model including interaction on both sex and care home residence status, but convergence of parameter estimates failed. The model with interaction on sex had the lowest AIC of all fitted models and, given also the relatively small number of care home residents in the dataset, we therefore focus on this model for interpretation and analysis.

Kaplan-Meier plots of mortality in relation to B.1.1.7 status are presented according to patient sex and age categories in Figure 2 (also provided separately for non-HOCI and HOCI inpatients in Figures S3-4, with HR estimates in Table S1).

**Figure 2.**
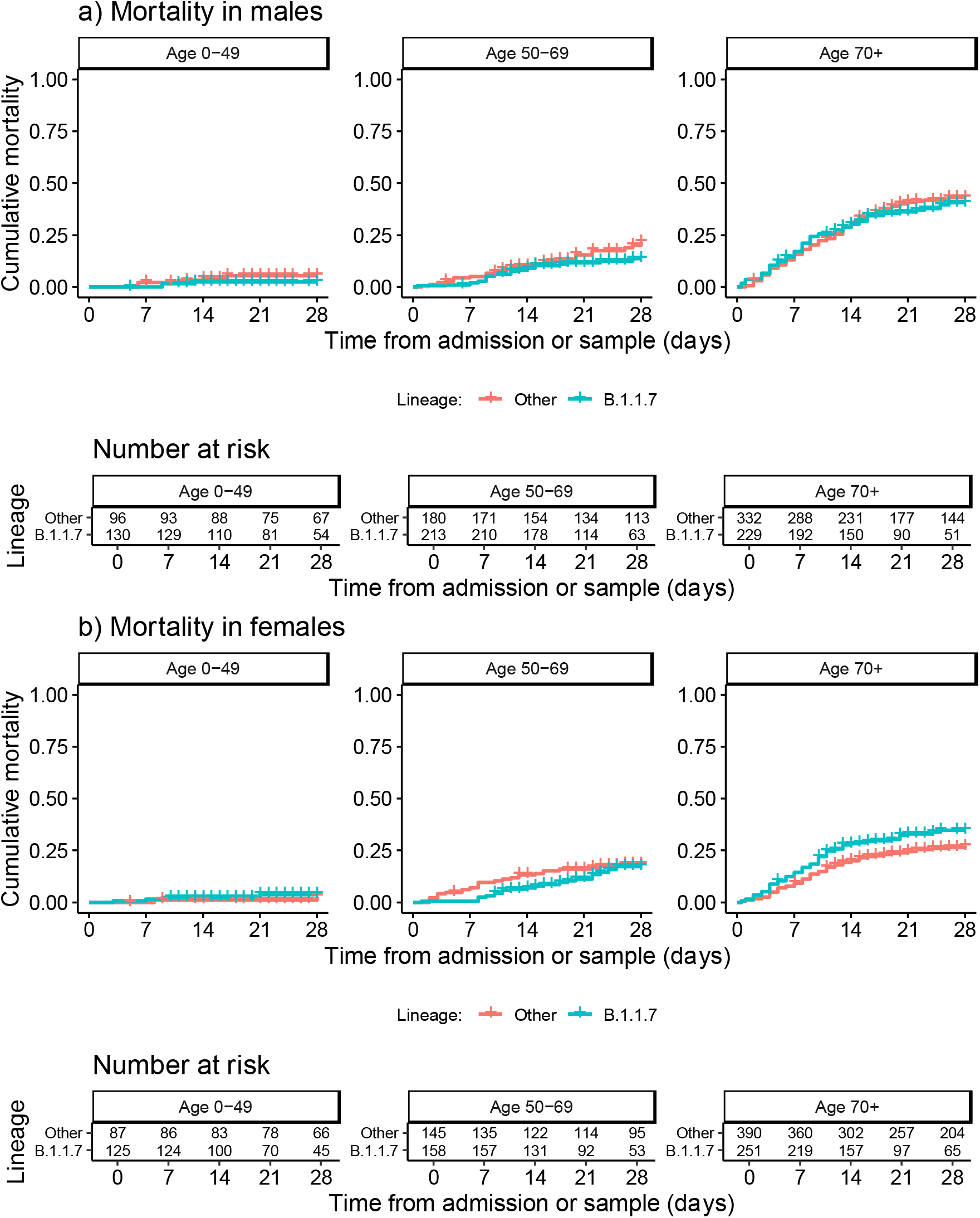
Kaplan-Meier plots of all-cause mortality among all inpatients in relation to lineage B.1.1.7 status, plotted according to patient sex and age categories. Date of sampling is used as the ‘zero’ time point for hospital-onset COVID-19 infections, with date of admission used for other patients. Naïve 95%CIs are plotted for illustrative purposes (these are not derived from the multilevel Cox models described).

### ITU admission outcome

Admission to ITU within 28 days was reported in 337 (14.4%) of 2341 inpatients (excluding 46 HOCI cases diagnosed after admission to ITU). On mixed effects multivariable Cox regression, the overall HR for ITU admission for lineage B.1.1.7 was 1.01 (95% CI 0.75-1.37, P=0.94) (Table 2). Within this model, male sex was a substantial risk factor for ITU admission (HR 1.33, 1.05-1.68; P=0.02). Age was also strongly associated with the risk of ITU admission, although the relationship estimated was non-linear with the greatest risk of this outcome at 65 years of age (Figure 1). The risk of ITU admission was higher in those with one or two significant comorbidities (HR 1.25 (0.92-1.71) for one comorbidity, 1.24 (0.89-1.74) for two and 0.79 (0.54-1.15) for ≥three *vs* none; P=0.03). Those with ethnicity other than White were estimated to be at higher risk of ITU admission, but ethnicity was not a statistically significant predictor evaluated over all categories (P=0.09). Pregnant women were found to be at lower risk of ITU admission (HR 0.13, 95% CI 0.02 to 0.98, P=0.048).

The addition of an interaction term between B.1.1.7 status and patient sex for the effect on ITU admission led to an improvement in model fit (P=0.0004 interaction test, P=0.002 lineage B.1.1.7 effects by sex vs no B.1.1.7 effect, LRTs). The estimated HR for ITU admission for lineage B.1.1.7 vs non-B.1.1.7 was 0.74 (95% CI 0.52-1.04) in male patients and 1.82 (95% CI 1.15-2.90) in female patients. There was no evidence for an interaction of B.1.1.7 status with patient age (P=0.11, LRT with 4 d.f.), ethnicity (P=0.74, LRT with 3 d.f.), comorbidity category (P=0.79, LRT with 3 d.f.), pregnancy (P=0.42, LRT with 1 d.f.) or care home residence (P=0.24, LRT with 1 d.f.) with ITU admission as the outcome. Kaplan-Meier plots of ITU admission in relation to B.1.1.7 status are presented according to patient sex and age categories in Figure 3 (also provided separately for non-HOCI and HOCI inpatients in Figures S5-6, with HR estimates in Table S1).

**Figure 3.**
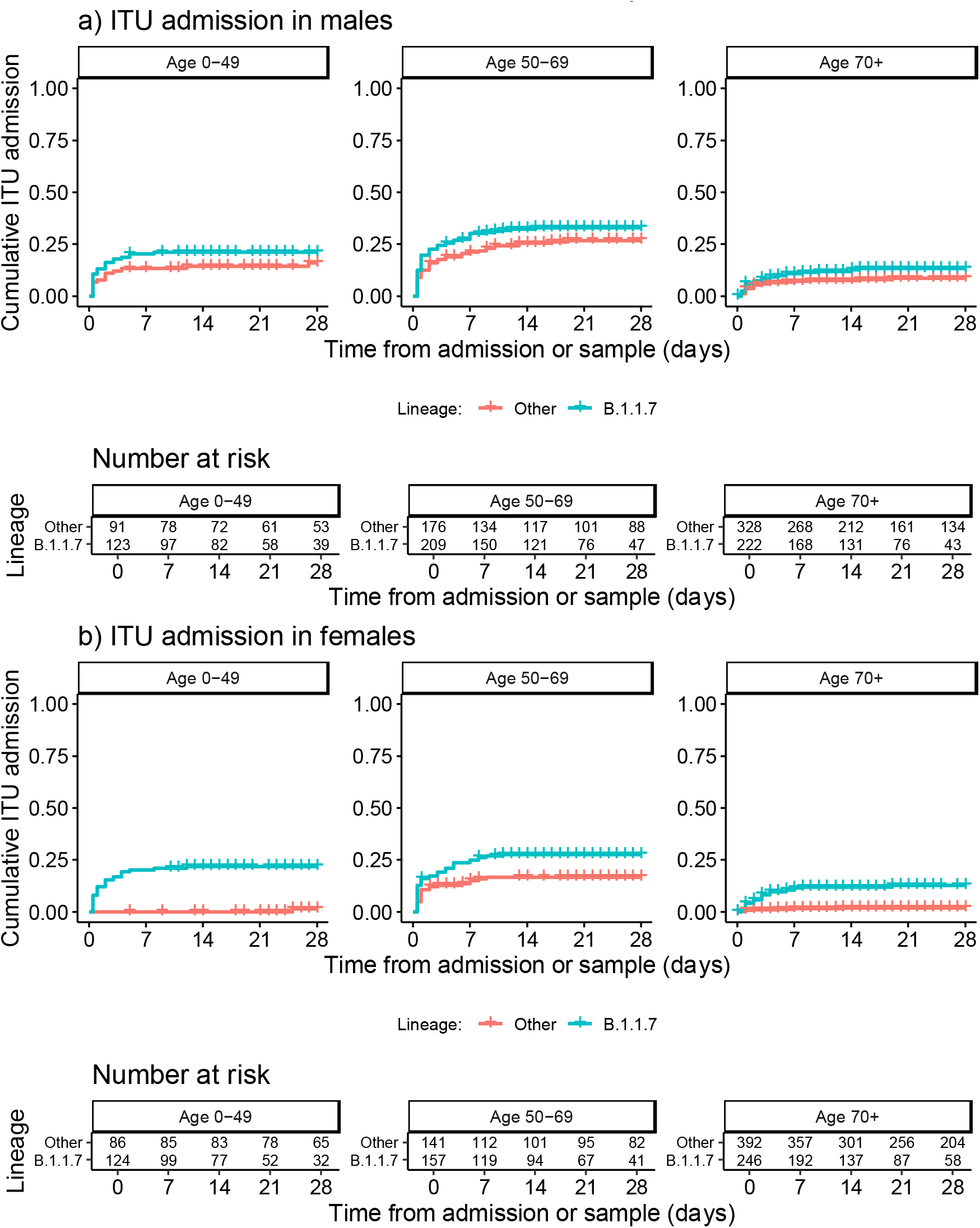
Kaplan-Meier plots of intensive therapy unit (ITU) admission among all inpatients in relation to lineage B.1.1.7 status, plotted according to patient sex and age categories. Date of sampling is used as the ‘zero’ time point for hospital-onset COVID-19 infections, with date of admission used for other patients. Naïve 95%CIs are plotted for illustrative purposes (these are not derived from the multilevel Cox models described).

## Discussion

Our findings provide the largest dataset on disease severity in hospitalized patients with lineage B.1.1.7 and the only one based on routine sequencing of all specimens from multiple hospitals. The overall hazard of mortality and ITU were unchanged for patients with lineage B.1.1.7 in comparison to other viral variants (HR 1.01, 95% CI 0.79-1.28; and 1.01, 95% CI 0.75-1.37, respectively).

These findings are in line with the results of a much smaller analysis of 341 (*n*=198 with B.1.1.7) hospital inpatients with viral sequencing over a similar time period in London, which found an adjusted mortality risk ratio for B.1.1.7 of 1.02 (95% CI 0.76-1.38)^19^. However, in contrast with this smaller study we also found evidence that B.1.1.7 infection appears to have a different impact on the disease course according to sex among hospitalised patients with SARS-CoV-2 infection, with increased hazard of both mortality and ITU admission associated with the variant for female but not male patients.

Several larger studies of disease severity in the UK have used PCR Spike (S) gene target failure (SGTF) as a surrogate for lineage B.1.1.7^20,21,22,23^. These studies, based on community testing data, all found evidence of an overall increased risk of mortality associated with lineage B.1.1.7, with reported hazard ratios of 1.64 (95% CI 1.32 to 2.04) by Challen et al.^20^, 1.55 (1.39 to 1.72) by Davies et al.^21^, 1.67 (1.34–2.09) by Grint et al.^22^ and 1.59 (1.25-2.03) by Patone et al.^23^. In the UK SGTF is only available as a marker for a subset of those patients who were first positive for SARS-CoV-2 on testing within the community; most people who die of COVID-19 were not previously tested within the community^20^ and the relevant PCR assay is not used by all laboratories, meaning that SGTF status is only available for 8.6% of deaths^21^. SGTF is an imperfect predictor of lineage B.1.1.7, and is much less accurate as a marker when prevalence of the variant is low (before mid-November 2020 in the UK)^24^.

The apparent overall differences in mortality risk observed in the SGTF analyses in comparison to our study do not necessarily represent inconsistent findings. Studies that are limited to patients who test positive in the community may be subject to selection biases linked to propensity to present for testing or rapidity of disease progression, whilst analyses that include only data from inpatients will not reflect the characteristics of the population as a whole. For example, increased disease severity may result in a higher proportion of subjects reaching the threshold for admission to hospital but not affect the mortality rate among those admitted to hospital. Our study also includes a subset of patients with probable nosocomial infection, whose characteristics and comorbidity profile differs greatly from the UK population as whole^25^.

Individuals testing positive in the community for an SGTF-associated variant had higher risk of hospitalisation, with OR of 1.58 (95% CI 1.50 to 1.67)^26^. This result was confirmed by a study of national health register-data from Denmark including 18,499 patients with viral genomes available in the period 1st January to 9th February 2021 which found an adjusted OR of 1.64 (95% CI, 1.32-2.04) for hospitalisation for B.1.1.7 compared with other lineages^27^. Taken together with the findings regarding mortality in the UK^20,21,22,23^, these results are consistent with an increased risk of mortality and hospitalisation among patients testing positive for B.1.1.7 in the community but no overall increase in mortality among the subset of patients admitted to hospital.

We found a significantly increased risk of both mortality (30%) and ITU admission (82%) in hospitalised female patients infected with B.1.1.7 but not in male patients. In contrast studies of community tested individuals found no interaction with sex for the effect of B.1.1.7 on mortality^21,23^, critical care admission^23^ or risk of hospitalisation^26^. However, these studies were all conducted among patients who first tested positive for SARS-CoV-2 within the community, and therefore they cannot rule out an interaction with sex for the impact of B.1.1.7 on disease severity among all people infected with the virus or among those admitted to hospital. Nationally collated data show that females accounted for 33.2% of patients admitted to ITU with COVID-19 in London, East and South East England between 1 September - 30 November 2020 rising to 36.2%, between 1 December 2020 to 21 January 2021 when lineage B.1.1.7 predominated^28^.

An impact of lineage B.1.1.7 on females that is not observed in males could potentially be explained by physiological differences. For example, increased ACE2 expression in females has been posited as one explanation for the relatively lower mortality and morbidity observed for COVID-19 for women in comparison to men ^29,30^. Lineage B.1.1.7 has mutations that increase binding of the viral spike protein to ACE2, thereby providing a plausible mechanism by which the new variant might have a differential effect on disease severity in males and females ^5,29,31^. Our results suggest a reduction in the risk of mortality or ITU admission associated with B.1.1.7 in comparison to other viral lineages among male inpatients, although this finding was not definitive with HR 95% CIs that included no effect for both outcomes.

Although ours is substantially the largest study of hospitalized patients with confirmed lineage B.1.1.7 and non-B.1.1.7 SARS-Cov-2 infection, it has a number of limitations. Primarily, whilst evaluation of disease severity among only hospital inpatients can give useful information on disease course and progression, analysis of only these patients cannot provide information on disease severity across all SARS-CoV-2 infections in the population as a whole. In addition, ITU admission can be difficult to interpret as a measure of disease severity among inpatients. For instance, admission to ITU may reflect the presence of severe disease but also local decisions around the benefit or lack thereof to frail patients, which may be influenced by bed numbers and availability of respiratory support in non-critical care settings. Our primary analysis also includes cases of hospital-acquired infection, but exclusion of these HOCI cases from our analyses yielded similar findings (Table S1).

A further limitation of our analysis is that we do not have any information on vaccination status for individual patients. Our dataset covers a period in which a national vaccination program was being initiated for HCWs and the elderly population in the UK, starting with those aged 80 years and above from 8th December 2020. This is a potential explanation for the observed protective interaction effect between care home residence and B.1.1.7 on mortality, as care home residents were prioritised for vaccination around the time that this viral variant was increasing in prevalence. Vaccine breakthrough infections are well described, particularly in partially vaccinated subjects^32^.

The findings may have implications for hospital practice and public health policy, both in the UK and in other countries where lineage B.1.1.7 is now dominant or spreading. Although lineage B.1.1.7 was not associated with an overall increase in mortality among hospitalized patients, our investigation suggests that lineage B.1.1.7 may be associated with higher ITU admission and death in females compared to non-B.1.1.7 within this group. The dominance of lineage B.1.1.7 in the UK precludes ongoing comparison with earlier non-B.1.1.7 variants, and there is now concern regarding the spread of other lineages in the UK and elsewhere^33^. There is a need for ongoing large scale sequencing of SARS-CoV-2 cases linked to data on patient characteristics and outcomes in order to generate timely information regarding the associations between viral lineages and disease severity.

## Supporting information

List of consortia members

## Data Availability

The sequence data analysed are included within publicly available datasets (https://www.cogconsortium.uk/data/). However, due to data governance restrictions it is not possible to openly share the associated patient characteristics and clinical outcome data for the analysis described, as these are considered sensitive and full anonymisation is not possible.

## Data Availability

Due to potential risk of de-identification of pseudonymized RNA sequencing data the raw data will be available under controlled access in the EGA repository, [will be added upon completion of peer review/ acceptance]. Count and metadata tables (patient-ID, sex, age, cell type, QC metrics per cell) can be found at FigShare: [will be added upon completion of peer review/ acceptance]. In addition, these data can be further visualized and analyzed in the Magellan COVID-19 data explorer at https://digital.bihealth.org [will be publicly available upon completion of peer review/ acceptance].

## Acknowledgments

This report was produced by members of the **COG-UK HOCI Variant substudy consortium. COG-UK HOCI** is part of **COG-UK**.

## Contributors

OTS, FATB, CV, JAGA, ACJ, NF, ACH and JB planned the analysis and drafted the first draft of the manuscript. AAM, AHB, TC, AdSF, SG, TK, RKS, BK, IRM, GM, MP, EP, PR, SR, JFT, SPW, EWD, PW, RW, TCM, AH, JH, TWM, GN, DGP, CFP, JRP, SCR, KS, TIdS, LBS, ECT, AAW extracted and provided sequencing data and patient characteristics and outcome data. OTS, FATB, CV and JAGA had full access to and verified the final collated dataset. FATB, CV and JAGA carried out phylogenetic lineage assignments and merged the final dataset for analysis, and OTS carried out statistical modelling. All authors reviewed the final manuscript and approved this for submission.

## References

1. Davies NG, Abbott S, Barnard RC, et al. Estimated transmissibility and severity of novel SARS-CoV-2 Variant of Concern 202012/01 in England. medRxiv 2021;2020.12.24.20248822.

2. Volz E, Mishra S, Chand M, et al. Transmission of SARS-CoV-2 Lineage B.1.1.7 in England: Insights from linking epidemiological and genetic data. medRxiv 2021;2020.12.30.20249034.

3. Lineage-specific growth of SARS-CoV-2 B.1.1.7 during the English national lockdown - SARS-CoV-2 coronavirus / nCoV-2019 Genomic Epidemiology [Internet]. Virological. 2020 [cited 2021 Feb 8];Available from: https://virological.org/t/lineage-specific-growth-of-sars-cov-2-b-1-1-7-during-the-english-national-lockdown/575

4. PANGO lineages [Internet]. [cited 2021 Feb 9];Available from: https://cov-lineages.org/global_report.html

5. Starr TN, Greaney AJ, Hilton SK, et al. Deep Mutational Scanning of SARS-CoV-2 Receptor Binding Domain Reveals Constraints on Folding and ACE2 Binding. Cell 2020;182(5):1295-1310.e20.

6. Kemp SA, Collier DA, Datir RP, et al. SARS-CoV-2 evolution during treatment of chronic infection. Nature 2021;1–10.

7. NERVTAG paper on COVID-19 variant of concern B.1.1.7 [Internet]. GOV.UK. [cited 2021 Feb 9];Available from: https://www.gov.uk/government/publications/nervtag-paper-on-covid-19-variant-of-concern-b117

8. COVID-19: the green book, chapter 14a [Internet]. GOV.UK. [cited 2021 May 21];Available from: https://www.gov.uk/government/publications/covid-19-the-green-book-chapter-14a

9. COVID-19: epidemiological definitions of outbreaks and clusters [Internet]. GOV.UK. [cited 2021 Feb 9];Available from: https://www.gov.uk/government/publications/covid-19-epidemiological-definitions-of-outbreaks-and-clusters

10. cov-lineages/pangolin [Internet]. CoV-lineages; 2021 [cited 2021 Feb 9]. Available from: https://github.com/cov-lineages/pangolin

11. Therneau T. coxme: Mixed Effects Cox Models. R package version 2.2-16. [Internet]. 2020. Available from: https://CRAN.R-project.org/package=coxme

12. Kahan BC, Rushton H, Morris TP, Daniel RM. A comparison of methods to adjust for continuous covariates in the analysis of randomised trials. BMC Medical Research Methodology 2016;16(1):42.

13. Bartlett JW, Harel O, Carpenter JR. Asymptotically Unbiased Estimation of Exposure Odds Ratios in Complete Records Logistic Regression. Am J Epidemiol 2015;182(8):730–6.

14. Hughes RA, Heron J, Sterne JAC, Tilling K. Accounting for missing data in statistical analyses: multiple imputation is not always the answer. Int J Epidemiol 2019;48(4):1294–304.

15. R Core Team. R: A language and environment for statistical computing. [Internet]. Vienna, Austria.: R Foundation for Statistical Computing; 2019 [cited 2019 Sep 1]. Available from: https://www.R-project.org/

16. Wickham H. ggplot2: Elegant Graphics for Data Analysis [Internet]. Springer-Verlag New York; 2016. Available from: https://ggplot2.tidyverse.org

17. Wickham H, Averick M, Bryan J, et al. Welcome to the Tidyverse. Journal of Open Source Software 2019;4(43):1686.

18. Kassambara A, Kosinski M, Biecek P, Fabian S. survminer: Drawing Survival Curves using “ggplot2” [Internet]. 2020 [cited 2021 Feb 11]. Available from: https://CRAN.R-project.org/package=survminer

19. Frampton D, Rampling T, Cross A, et al. Genomic characteristics and clinical effect of the emergent SARS-CoV-2 B.1.1.7 lineage in London, UK: a whole-genome sequencing and hospital-based cohort study. The Lancet Infectious Diseases [Internet] 2021 [cited 2021 Apr 27];0(0). Available from: https://www.thelancet.com/journals/laninf/article/PIIS1473-3099(21)00170-5/abstract

20. Challen R, Brooks-Pollock E, Read JM, Dyson L, Tsaneva-Atanasova K, Danon L. Risk of mortality in patients infected with SARS-CoV-2 variant of concern 202012/1: matched cohort study. BMJ 2021;372:n579.

21. Davies NG, Jarvis CI, Edmunds WJ, Jewell NP, Diaz-Ordaz K, Keogh RH. Increased mortality in community-tested cases of SARS-CoV-2 lineage B.1.1.7. Nature 2021;1–5.

22. Grint DJ, Wing K, Williamson E, et al. Case fatality risk of the SARS-CoV-2 variant of concern B.1.1.7 in England, 16 November to 5 February. Eurosurveillance 2021;26(11):2100256.

23. Patone M, Thomas K, Hatch R, et al. Analysis of severe outcomes associated with the SARS-CoV-2 Variant of Concern 202012/01 in England using ICNARC Case Mix Programme and QResearch databases. medRxiv 2021;2021.03.11.21253364.

24. Guerra-Assunção JA, Randell PA, Boshier FAT, et al. Reliability of Spike Gene Target Failure for ascertaining SARS-CoV-2 lineage B.1.1.7 prevalence in a hospital setting. medRxiv 2021;2021.04.12.21255084.

25. Bhattacharya A, Collin SM, Stimson J, et al. Healthcare-associated COVID-19 in England: a national data linkage study. medRxiv 2021;2021.02.16.21251625.

26. Nyberg T, Twohig KA, Harris RJ, et al. Increased risk of hospitalisation for COVID-19 patients infected with SARS-CoV-2 variant B.1.1.7. arXiv:210405560 [stat] [Internet] 2021 [cited 2021 Apr 28];Available from: http://arxiv.org/abs/2104.05560

27. Bager P, Wohlfahrt J, Fonager J, et al. Increased Risk of Hospitalisation Associated with Infection with SARS-CoV-2 Lineage B.1.1.7 in Denmark [Internet]. Rochester, NY: Social Science Research Network; 2021 [cited 2021 Mar 19]. Available from: https://papers.ssrn.com/abstract=3792894

28. ICNARC – Reports [Internet]. 2021 [cited 2021 Feb 10];Available from: https://www.icnarc.org/Our-Audit/Audits/Cmp/Reports

29. Gagliardi MC, Tieri P, Ortona E, Ruggieri A. ACE2 expression and sex disparity in COVID-19. Cell Death Discovery 2020;6(1):1–2.

30. Foresta C, Rocca MS, Di Nisio A. Gender susceptibility to COVID-19: a review of the putative role of sex hormones and X chromosome. J Endocrinol Invest [Internet] 2020 [cited 2021 Feb 9];Available from: https://doi.org/10.1007/s40618-020-01383-6

31. Santos JC, Passos GA. The high infectivity of SARS-CoV-2 B.1.1.7 is associated with increased interaction force between Spike-ACE2 caused by the viral N501Y mutation. bioRxiv 2021;2020.12.29.424708.

32. Abu-Raddad LJ, Chemaitelly H, Butt AA. Effectiveness of the BNT162b2 Covid-19 Vaccine against the B.1.1.7 and B.1.351 Variants. New England Journal of Medicine 2021;0(0):null.

33. Thiagarajan_2021_Why is India having a covid-19 surge.pdf [Internet]. [cited 2021 May 17];Available from: https://www.bmj.com/content/bmj/373/bmj.n1124.full.pdf

