## Supplementary Figures and Tables for "SARS-CoV-2 lineage B.1.1.7 is associated with greater disease severity among hospitalised women but not men"

**Supplementary Table 1** Hazard ratios for the outcomes of mortality and intensive therapy unit admission associated with lineage B.1.1.7 from mixed effects Cox models, with sensitivity analyses limited to hospital-onset COVID-19 infection (HOCI) and non-HOCI cases.

| | | **Interaction by sex** | |
|---|---|---|---|
| | **Overall effect of lineage B.1.1.7** | **Effect of lineage B.1.1.7 in males** | **Effect of lineage B.1.1.7 in females** |
| *Mortality* | | | |
| All inpatients | 1.01 (0.79 to 1.28) | 0.82 (0.61 to 1.10) | 1.30 (0.95 to 1.78) |
| Excluding HOCI cases | 1.2 (0.86 to 1.69) | 0.94 (0.63 to 1.4) | 1.64 (1.06 to 2.52) |
| Only HOCI cases | 0.83 (0.55 to 1.23) | 0.72 (0.43 to 1.19) | 0.97 (0.57 to 1.63) |
| *ITU admission* | | | |
| All inpatients | 1.01 (0.75 to 1.37) | 0.74 (0.52 to 1.04) | 1.82 (1.15 to 2.90) |
| Excluding HOCI cases | 0.88 (0.62 to 1.24) | 0.66 (0.45 to 0.97) | 1.53 (0.92 to 2.57 |
| Only HOCI cases | 0.96 (0.42 to 2.18) | 0.74 (0.27 to 2.02) | 1.42 (0.42 to 4.77) |

Results shown as hazard ratio (95% CI).

**Table S2** Proportion of SARS-CoV-2 due to lineage B.1.1.7 for all male inpatient sequenced samples according to patient characteristics

| | **Lineage B.1.1.7 (n=573)** | **Not lineage B.1.1.7 (n=610)** | **Total (n=1183)** |
|---|---|---|---|
| **Age Group** | | | |
| 0-11 | 12 (75) | 4 (25) | 16 (100) |
| 12-24 | 9 (52.9) | 8 (47.1) | 17 (100) |
| 25-34 | 27 (65.9) | 14 (34.1) | 41 (100) |
| 35-49 | 82 (53.9) | 70 (46.1) | 152 (100) |
| 50-69 | 213 (54.1) | 181 (45.9) | 394 (100) |
| 70-79 | 107 (41.8) | 149 (58.2) | 256 (100) |
| 80+ | 123 (40.1) | 184 (59.9) | 307 (100) |
| **Sex** | | | |
| Female | 0 (NaN) | 0 (NaN) | 0 (NaN) |
| Male | 573 (48.4) | 610 (51.6) | 1183 (100) |
| **Sample week starting:** | | | |
| 16/11/2020 | 9 (10.5) | 77 (89.5) | 86 (100) |
| 23/11/2020 | 12 (10.3) | 105 (89.7) | 117 (100) |
| 30/11/2020 | 31 (24.2) | 97 (75.8) | 128 (100) |
| 07/12/2020 | 23 (26.4) | 64 (73.6) | 87 (100) |
| 14/12/2020 | 71 (45.5) | 85 (54.5) | 156 (100) |
| 21/12/2020 | 117 (57.4) | 87 (42.6) | 204 (100) |
| 28/12/2020 | 197 (75.5) | 64 (24.5) | 261 (100) |
| 04/01/2021 | 113 (78.5) | 31 (21.5) | 144 (100) |
| **Patient Class** | | | |
| HCW | 1 (33.3) | 2 (66.7) | 3 (100) |
| CAI* | 439 (55.6) | 351 (44.4) | 790 (100) |
| Indeterminate HCAI† | 29 (28.7) | 72 (71.3) | 101 (100) |
| Probable/definite HCAI‡ | 104 (36) | 185 (64) | 289 (100) |
| **Region** | | | |
| Glasgow | 40 (30.5) | 91 (69.5) | 131 (100) |
| Hampshire | 39 (57.4) | 29 (42.6) | 68 (100) |
| London | 459 (64.3) | 255 (35.7) | 714 (100) |
| South Yorkshire | 35 (13) | 235 (87) | 270 (100) |
| **Ethnicity** | | | |
| White | 244 (37.7) | 403 (62.3) | 647 (100) |
| Black | 101 (54) | 86 (46) | 187 (100) |
| Asian | 62 (72.1) | 24 (27.9) | 86 (100) |
| Mixed or other | 105 (68.6) | 48 (31.4) | 153 (100) |
| Unknown | 61 (55.5) | 49 (44.5) | 110 (100) |
| **Patient characteristics** | | | |
| Obese (BMI>=35) | 47 (52.2) [N=573] | 43 (47.8) [N=610] | 90 (100) [N=1183] |
| Pregnant | NA | NA | NA |
| Care home resident | 21 (41.2) [N=573] | 30 (58.8) [N=609] | 51 (100) [N=1182] |

| | | | |
|---|---|---|---|
| **Comorbidities** | | | |
| None | 181 (58) | 131 (42) | 312 (100) |
| One | 153 (48) | 166 (52) | 319 (100) |
| Two | 129 (46.9) | 146 (53.1) | 275 (100) |
| Three or more | 110 (39.7) | 167 (60.3) | 277 (100) |
| Not recorded | 0 (NA) | 0 (NA) | 0 (NA) |
| **Died within 28d** | 112 (38.4) [N=572] | 180 (61.2) [N=608] | 292 (100) [N=1180] |
| **Admitted to ITU within 28d⁋** | 121 (58.5) [N=554] | 86 (41.5) [N=595] | 207 (100) [N=1149] |

Data shown as *n* (%), with [N] with available data shown where missing values possible. *Diagnosed at or ≤2 days from admission. †Diagnosed 3-7 days from admission. ‡Diagnosed ≥8 days from admission. ⁋Excluding patients admitted to ITU prior to SARS-CoV-2 diagnosis. CAI, community-acquired infection; HCAI, healthcare-associated infection; HCW, healthcare worker; ITU, intensive therapy unit.

**Table S3** Proportion of SARS-CoV-2 due to lineage B.1.1.7 for all female inpatient sequenced samples according to patient characteristics

| | **Lineage B.1.1.7 (n=534)** | **Not lineage B.1.1.7 (n=624)** | **Total (n=1158)** |
|---|---|---|---|
| **Age Group** | | | |
| 0-11 | 3 (37.5) | 5 (62.5) | 8 (100) |
| 12-24 | 11 (64.7) | 6 (35.3) | 17 (100) |
| 25-34 | 34 (60.7) | 22 (39.3) | 56 (100) |
| 35-49 | 77 (58.8) | 54 (41.2) | 131 (100) |
| 50-69 | 158 (52.1) | 145 (47.9) | 303 (100) |
| 70-79 | 101 (42.6) | 136 (57.4) | 237 (100) |
| 80+ | 150 (36.9) | 256 (63.1) | 406 (100) |
| **Sex** | | | |
| Female | 534 (46.1) | 624 (53.9) | 1158 (100) |
| Male | 0 (NaN) | 0 (NaN) | 0 (NaN) |
| **Sample week starting:** | | | |
| 16/11/2020 | 6 (6.5) | 87 (93.5) | 93 (100) |
| 23/11/2020 | 14 (13.1) | 93 (86.9) | 107 (100) |
| 30/11/2020 | 28 (24.1) | 88 (75.9) | 116 (100) |
| 07/12/2020 | 32 (27.1) | 86 (72.9) | 118 (100) |
| 14/12/2020 | 67 (42.1) | 92 (57.9) | 159 (100) |
| 21/12/2020 | 103 (51.8) | 96 (48.2) | 199 (100) |
| 28/12/2020 | 164 (74.9) | 55 (25.1) | 219 (100) |
| 04/01/2021 | 120 (81.6) | 27 (18.4) | 147 (100) |
| **Patient Class** | | | |
| HCW | 6 (37.5) | 10 (62.5) | 16 (100) |
| CAI* | 408 (54.7) | 338 (45.3) | 746 (100) |
| Indeterminate HCAI† | 25 (22.3) | 87 (77.7) | 112 (100) |
| Probable/definite HCAI‡ | 95 (33.5) | 189 (66.5) | 284 (100) |
| **Region** | | | |
| Glasgow | 51 (32.5) | 106 (67.5) | 157 (100) |
| Hampshire | 35 (63.6) | 20 (36.4) | 55 (100) |
| London | 412 (67.3) | 200 (32.7) | 612 (100) |
| South Yorkshire | 36 (10.8) | 298 (89.2) | 334 (100) |
| **Ethnicity** | | | |
| White | 296 (41) | 426 (59) | 722 (100) |
| Black | 73 (52.5) | 66 (47.5) | 139 (100) |
| Asian | 56 (55.4) | 45 (44.6) | 101 (100) |
| Mixed or other | 81 (65.3) | 43 (34.7) | 124 (100) |
| Unknown | 28 (38.9) | 44 (61.1) | 72 (100) |
| **Patient characteristics** | | | |
| Obese (BMI>=35) | 75 (50.3) [N=534] | 74 (49.7) [N=624] | 149 (100) [N=1158] |
| Pregnant | 25 (55.6) [N=530] | 20 (44.4) [N=624] | 45 (100) [N=1154] |
| Care home resident | 24 (32.9) [N=534] | 49 (67.1) [N=623] | 73 (100) [N=1157] |

| | | | |
|---|---|---|---|
| **Comorbidities** | | | |
| None | 156 (55.1) | 127 (44.9) | 283 (100) |
| One | 154 (46.7) | 176 (53.3) | 330 (100) |
| Two | 132 (44.9) | 162 (55.1) | 294 (100) |
| Three or more | 92 (36.9) | 157 (63.1) | 249 (100) |
| Not recorded | 0 (NA) | 2 (100) | 2 (100) |
| **Died within 28d** | 105 (44.7) [N=534] | 130 (55.3) [N=622] | 235 (100) [N=1156] |
| **Admitted to ITU within 28d⁋** | 99 (76.2) [N=527] | 31 (23.8) [N=619] | 130 (100) [N=1146] |

Data shown as *n* (%), with [N] with available data shown where missing values possible. *Diagnosed at or ≤2 days from admission. †Diagnosed 3-7 days from admission. ‡Diagnosed ≥8 days from admission. ⁋Excluding patients admitted to ITU prior to SARS-CoV-2 diagnosis. CAI, community-acquired infection; HCAI, healthcare-associated infection; HCW, healthcare worker; ITU, intensive therapy unit.

**Figure S1** Bar plot of the proportion of inpatient samples sent for sequencing that sequenced successfully for 6/8 hospitals from which data are available. The proportion of successfully sequenced samples was not different between weeks (chi-squared, p = 0.97).

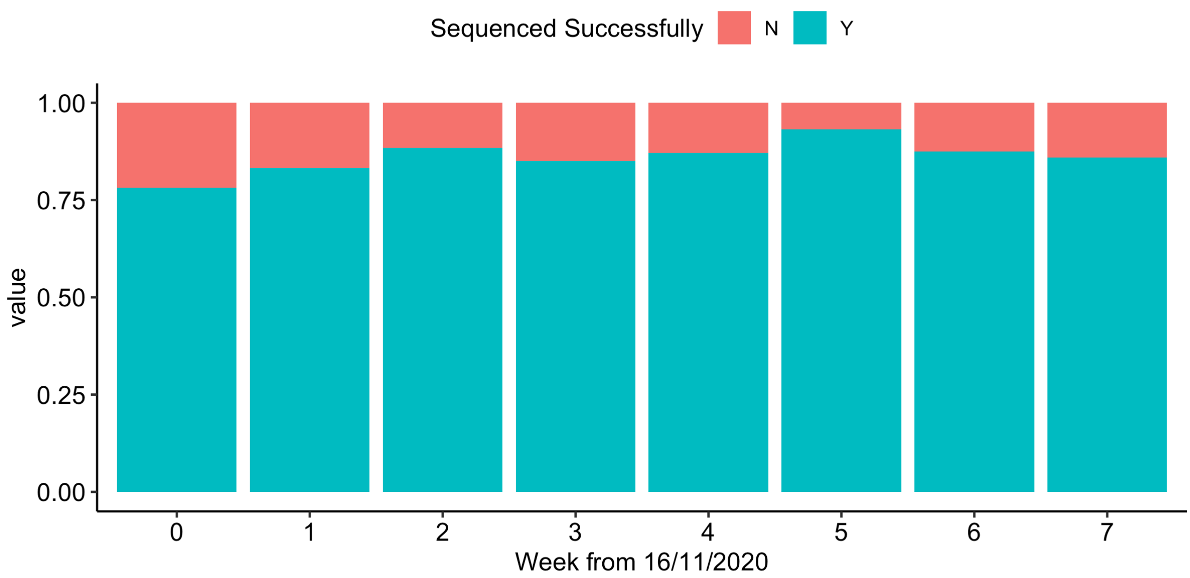

**Figure S2** Sequence counts by region, week of sample and patient type.

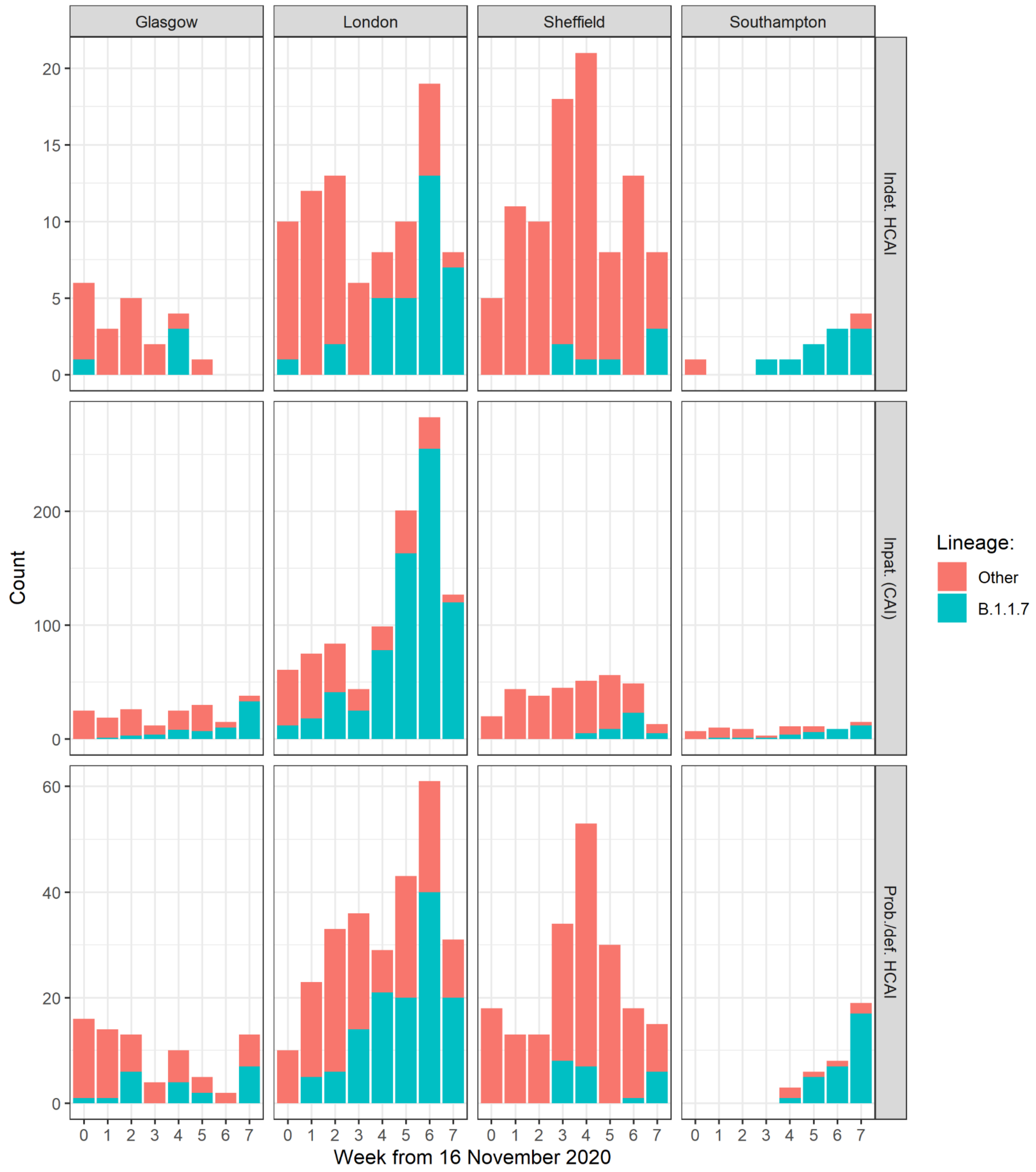

CAI, community-acquired infection; HCAI, healthcare-associated infection.

**Figure S3** Kaplan-Meier plots of all-cause mortality among all inpatients admitted with SARS-CoV-2 in relation to lineage B.1.1.7 status, plotted according to patient sex and age categories. Naïve 95%CIs are plotted for illustrative purposes (these are not derived from the multilevel Cox models described).

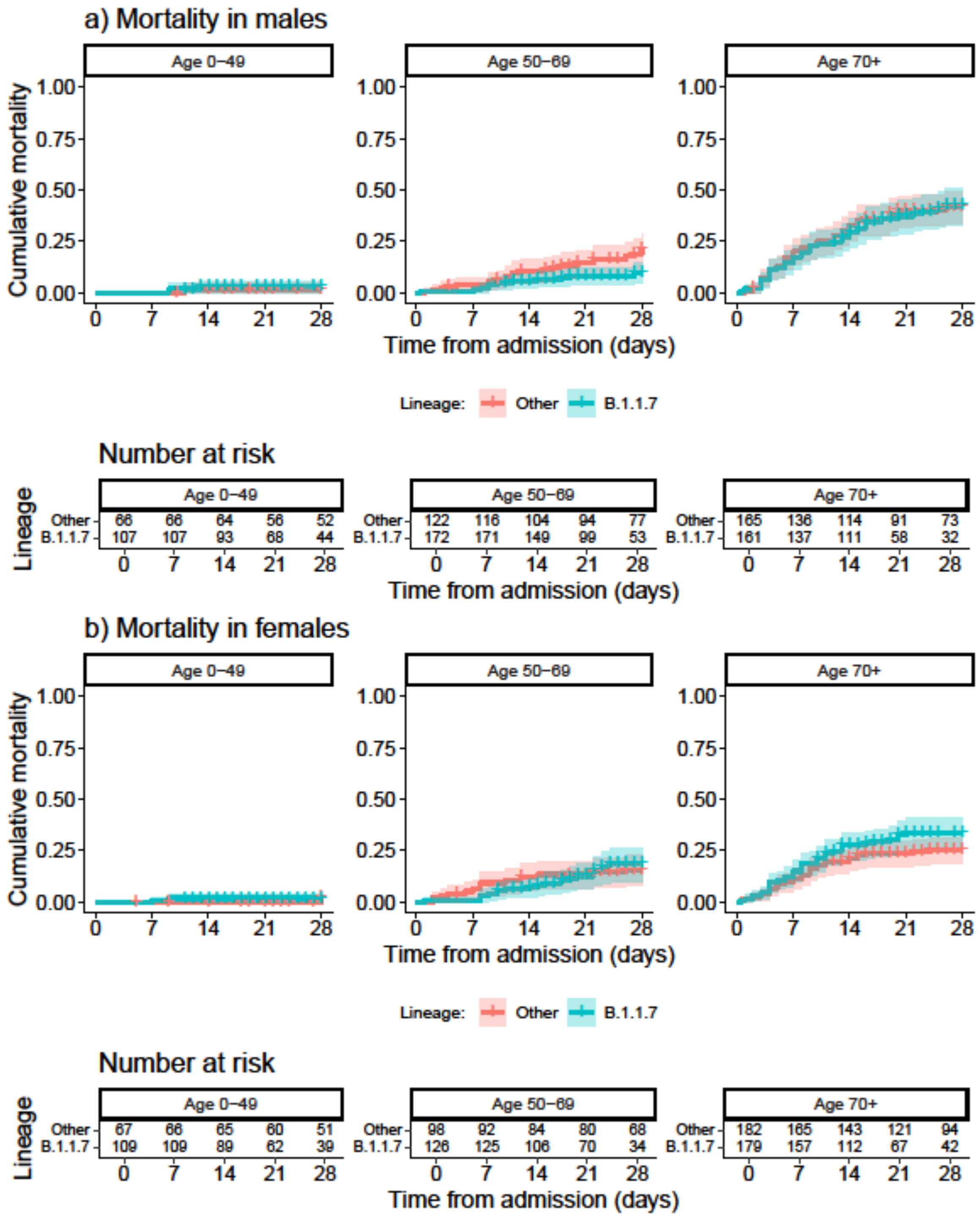

**Figure S4** Kaplan-Meier plots of all-cause mortality among all hospital-onset COVID-19 infection cases in relation to lineage B.1.1.7 status, plotted according to patient sex and age categories. Naïve 95%CIs are plotted for illustrative purposes (these are not derived from the multilevel Cox models described).

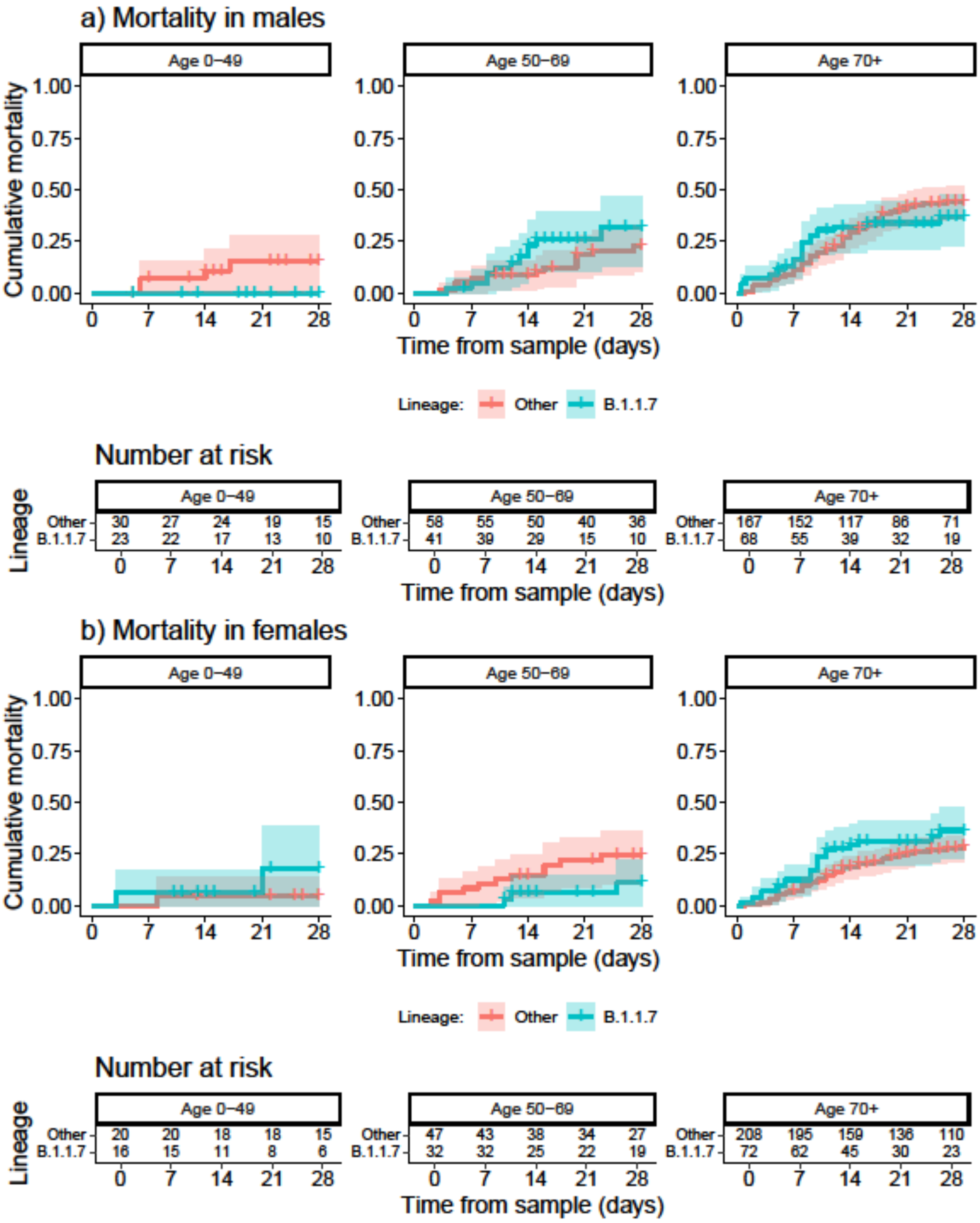

**Figure S5** Kaplan-Meier plots of intensive therapy unit (ITU) admission among all inpatients admitted with SARS-CoV-2 in relation to lineage B.1.1.7 status, plotted according to patient sex and age categories. Naïve 95%CIs are plotted for illustrative purposes (these are not derived from the multilevel Cox models described).

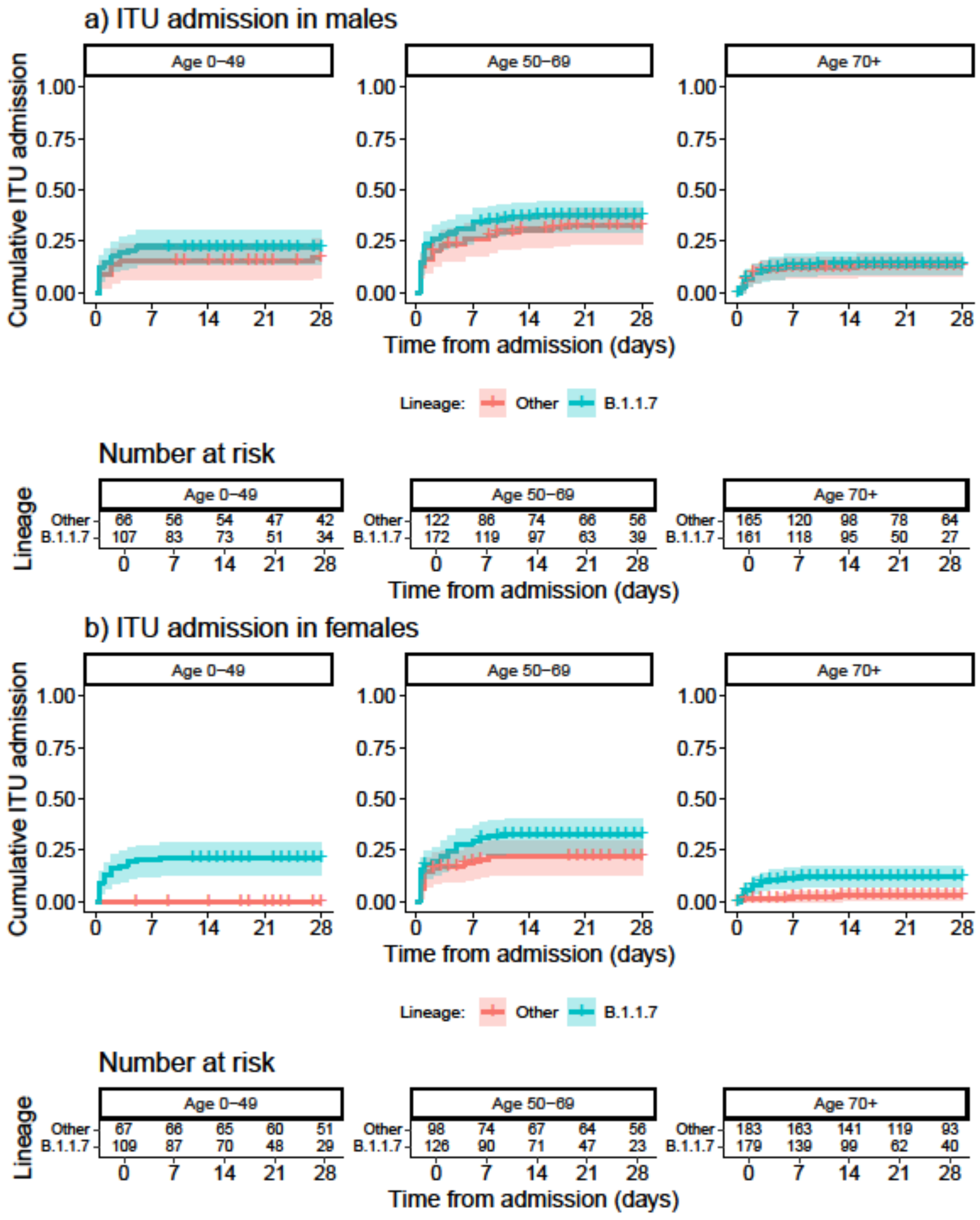

**Figure S6** Kaplan-Meier plots of intensive therapy unit (ITU) admission among all hospital-onset COVID-19 infection cases in relation to lineage B.1.1.7 status, plotted according to patient sex and age categories. Naïve 95%CIs are plotted for illustrative purposes (these are not derived from the multilevel Cox models described).

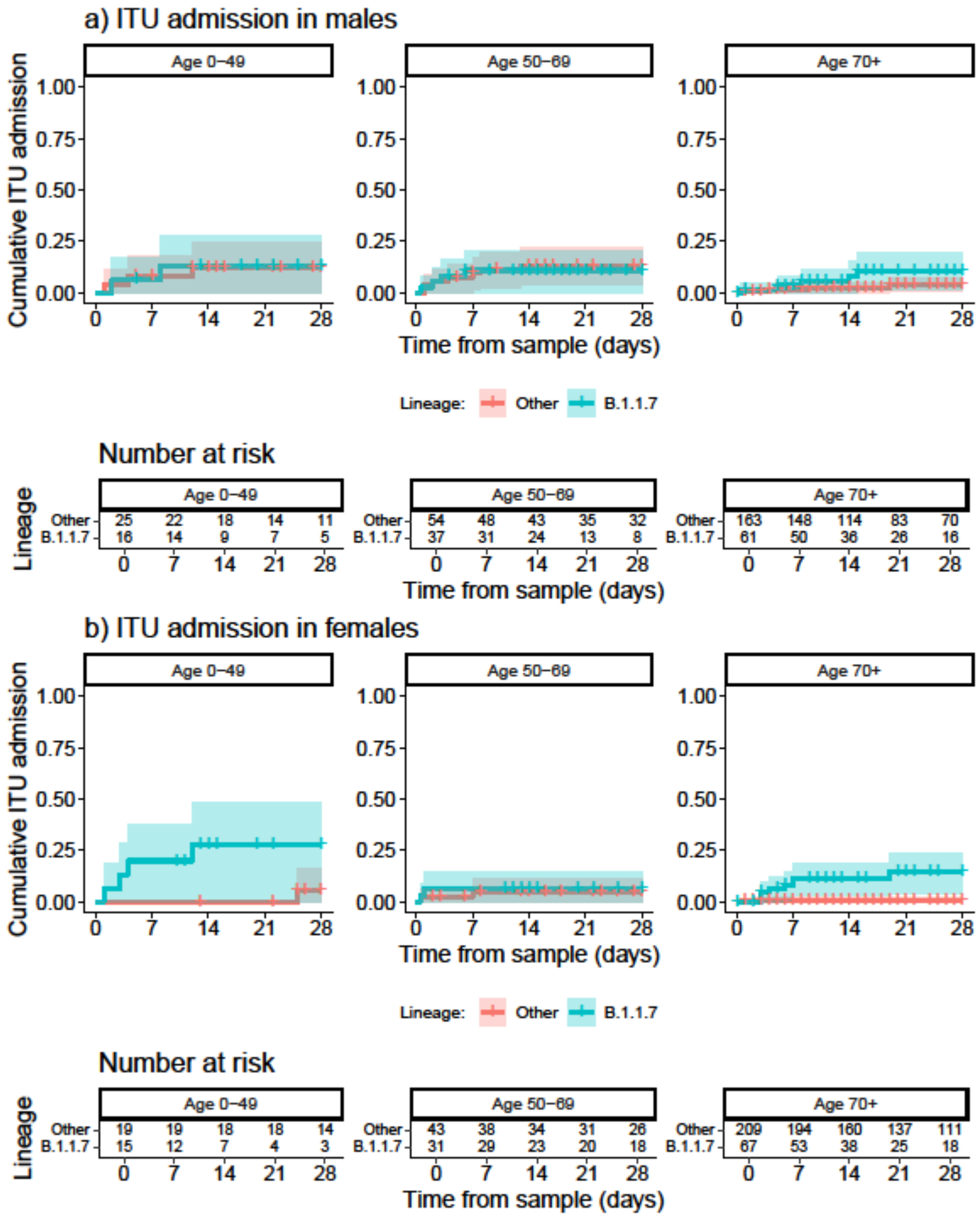
