## Supplementary material for "SARS-CoV-2 lineage B.1.1.7 is associated with greater disease severity among hospitalised women but not men": List of consortia members

### Data analysis and report writing

*Oliver Stirrup
*Florencia Tettamanti Boshier
*Cristina Venturini
*José Afonso Guerra-Assunção
Nick Freemantle
Andrew Copas
Andrew Hayward
Judith Breuer
*contributed equally

### Study oversight and quality assurance

**UCL Comprehensive Clinical Trials Unit**
James Blackstone
Leanne Hockey
Georgia Marley

### Collection of samples, sequence data and meta-data

**Barts Health Trust**
Teresa Cutino-Moguel
Tabassum Khan
Raghavendran Kulasegaran-Shylini
Beatrix Kele
David Harrington
Anna Riddell

**MRC-University of Glasgow Centre for Virus Research (CVR)**
Emma Thomson
Joseph Hughes
Ana Filipe

**Queen Elizabeth University Hospital, NHS Greater Glasgow & Clyde (QEUH)**
Guy Mollett
Christine Peters
Emma Thomson

**Guy's and St Thomas' NHS Foundation Trust and Centre for Clinical Infection & Diagnostics Research, King's College London**

Gaia Nebbia
Luke Blagdon Snell
Flavia Flaviani
Themoula Charalampous
Adela Alcolea-Medina
Bindi Patel
Tom G S Williams
Rahul Batra
Jonathan D Edgeworth

**Imperial College Healthcare NHS Trust**
James R Price
Alison H Holmes

**North West London Pathology**
Paul Randell
Pinglawathee Madona
Alison Cox

**Royal Free London NHS Foundation Trust**
Tabitha Mahungu
Sophie Weller
Jennifer Hart
Tanzina Haque
Dianne Irish

**UCL Pathogen Genomics Unit**
Judith Breuer
Rachel Williams
José Afonso Guerra-Assunção
Juanita Pang
Sunando Roy
Charlotte Williams
Helena Tutill
Nadua Bayzid
Marius Cotic

**University of Sheffield and Sheffield Teaching Hospitals NHS Foundation Trust**
Thushan de Silva

David Partridge
Matthew Parker
Luke Green
Benjamin Lindsey
Amy State
Alison Cope
Katie Johnson
Adrienn Angyal
Peijun Zhang
Max Whiteley
Marta Gallis Ramalho
Stella Christou
Stavroula Louka
Hailey Hornsby
Benjamin Foulkes
Paige Wolverson
Joe Heffer
Nikki Smith

**University of Portsmouth**
Samuel Robson
Angela Beckett
Salman Goudarzi
Chris Fearn
Kate Cook
Katie Loveson

**Portsmouth Hospitals University NHS Trust**
Sharon Glaysher
Scott Elliott

**University Hospital Southampton NHS Foundation Trust**
Kordo Saeed
Eleri Wilson-Davies
Adhyana Mahamana
Buddhini Samaraweera
Siona Silveira
Stephen Aplin
Sarah Jeremiah
Helen Umpleby

Helen Wheeler
Matthew Harvey
Thea Sass
Jacqui Prieto
Emanuela Pelosi

**St Georges University and Healthcare Trust**
Kenneth Laing
Adam Witney
Irene Monahan
Ngee Keong Tan
Joshua Taylor
Cassie Pope
Claudia Cardoso Pereira

**University College London Hospitals NHS foundation Trust**
**UCLH Advanced Pathogen Diagnostics Unit**
Eleni Nastouli
Catherine F Houlihan
Dan Frampton
Tommy Rampling
Matt Byott
Judith Heaney
Gee Yen Shin
Moira Spyer
Malin Bergstrom
Emilie Sanchez
Stavroula M Paraskevopoulou
Marios Margaritis
